# Current epidemiological situation of COVID-19 in the Republic of Belarus: characteristics of the epidemic process, sanitary and anti-epidemic measures

**DOI:** 10.1101/2022.03.10.22271815

**Authors:** Ala M Dashkevich, Natalia D Kolomiets, Veronika S Vysotskaya, Iryna N Hlinskaya, Anzhela L Skuranovich, Aliaksandr A Tarasenka, Inna A Karaban, Elena L Gasich

## Abstract

This is an analysis of the features of the COVID-19 pandemic the population of the Republic of Belarus from February 2020 to January 2022, the characteristics of sanitary and anti-epidemic measures carried out in the country, assessment of study the safety (tolerance) of the vaccines used the epidemiological efficacy of the vaccination.

A retrospective analysis of COVID-19 cases in the Republic of Belarus from the beginning of registration (February 28, 2020) to January, 3, 2022 was performed. Vaccine safety (tolerance) and efficacy were assessed in an observational study. Safety (tolerance) was assessed by presence/absence of adverse reactions: general (fever, malaise, headache, muscle pain, runny nose, nausea, vomiting, sore throat, etc) and local ones (redness, swelling, soreness at the injection place).

The COVID-19 pandemic in the Republic of Belarus is characterized by successive development stages: from no cases in early 2020 to detected cases where most individuals had no history of contact with COVID-19 patients; periods of rising and falling incidence

Vaccines against COVID-19 (Gam-COVID-Vac (Russia), inactivated vaccine against SARS-CoV-2 (Vero Cell) Sinopharm / BIBP (China) demonstrated a high safety profile in mass vaccination of the population of the Republic of Belarus.

## 1. Introduction

In December 2019, there were reports of an outbreak of a new infection in Wuhan (China), the etiological factor of which was a new strain of β coronavirus, subsequently called SARS-CoV-2 [1]. In a short time, the infection actively spread throughout the countries and on March 11, 2020, the World Health Organization characterized the developing situation as a pandemic [2].

As of January 3, 2022 a total more than 290 million cases and more than 5 million deaths were registered worldwide according to public sources [3].

In the emerging epidemiological situation, vaccination is an important aspect in countering COVID-19.

To study the COVID-19 pandemic features among the population of the Republic of Belarus from February 2020 to January 2021, characterize the sanitary and anti-epidemic measures carried out in the country, assess the safety (tolerance) of the vaccines used and the epidemiological efficacy of the vaccination.

## 2. Materials and Methods

A retrospective analysis of COVID-19 cases in the Republic of Belarus from the beginning of registration (February 28, 2020) to January, 3, 2022 was performed. To assess the COVID-19 case detection dynamics, official registration data available on the website of the Ministry of Health of the Republic of Belarus were used.

The population of the Republic of Belarus in 2021 was 9 349 645 [4].

Coefficients characterizing the epidemic were considered.

The infection reproduction index (R, baseline reproductive number) was calculated based on the number of detected COVID-19 cases over the past 7 days. In general, R > 1 indicates the presence (persistence) of the potential of an infectious disease for epidemic spread in a particular population; R < 1 indicates the absence of such potential [5].

The Sputnik V vaccine is a combined vector 2-component vaccine consisting of a recombinant human adenovirus type 26 (rAd26) vector and a recombinant human adenovirus type 5 (rAd5) vector, both components carrying the SARS-CoV-2 full-length protein S gene [6, 7]. Each vaccine dose contains (1.0±0.5)×10^11^ per dose of each of the recombinant adenoviruses; 0.5 ml per dose for intramuscular injection [8].

Vaccine against SARS-CoV-2 (Vero Cell) Sinopharm / BIBP (PRC) -each vaccine dose (0.5 ml) contains 3.9-10.4 units of inactivated antigen SARS-CoV-2 [9].

Vaccine safety (tolerance) and efficacy were assessed in an observational study. Safety (tolerance) was assessed by presence/absence of adverse reactions: general (fever, malaise, headache, muscle pain, runny nose, nausea, vomiting, sore throat, etc) and local ones (redness, swelling, soreness at the injection place).

The efficacy rate (E) was calculated according to the formula: E(%)=100*(b-a)/b, where

- E is the efficacy rate,
- a is COVID-19 incidence rate (morbidity) among vaccinated individuals,
- b is COVID-19 incidence rate (morbidity) among unvaccinated individuals

The epidemiological efficacy index (K) was calculated according to the formula: K=b/a, where “a” is disease incidence among vaccinated people, “b” is disease incidence among unvaccinated ones [10].

The data were processed using the Excel 2010 statistical software suite. Confidence intervals (95% CI) were calculated using the method of A.Wald, J.Wolfowitz, corrected by A.Agresti, B.A.Coull [11].

Sanger sequencing was performed according to the protocol published December 26th, 2020 Geneva [Protocol for specific RT-PCRs for marker regions of the Spike region indicative of the UK SARS-CoV2 variant B.1.1.7 and the South African variant 501Y.V2].

To prepare DNA libraries for sequencing, we used the Ligation Sequencing kit 1D (SQK-LSK109) from Oxford Nanopore Technologies (Great Britain) and NEBNext FFPE Repair Mix (M6630) and NEBNext End repair / dA-tailing Module reagents. (E7546), NEBNext Quick Ligation Module (E6056) from New England Biolabs (USA), and AMPure XP magnetic particles from Beckman Coulter Life Sciences (Germany).

Sequencing was performed on a MinION instrument (Oxford Nanopore Technologies, UK). The analysis of the obtained data was carried out using the kromsatel bioinformatic software [https://github.com/masikol/kromsatel] – a tool for separating “raw” reads into non-chimeric fragments in accordance with the primer scheme described in the ARTIC V3 protocol; Bowtie2 is a program for aligning the obtained nucleotide sequences with respect to the reference file; samtools and bcftools are software packages for processing alignment files and building a consensus sequence. [nCoV-2019 sequencing protocol v3 (LoCost) V.3].

## 3. Results

### 3.1. Characteristics of the epidemic process of COVID-19

During the analyzed period, the epidemic process of COVID-19 in the Republic of Belarus has passed several stages of development (the absence of COVID-19 cases in the country (until 28.02.2020); registration of individual infection cases that came from abroad followed by local pathogen spread among the country’s population (March 2020); a local spread of COVID-19 among individuals who had contact with infected people (since April 2020); the detection of cases where patients had no history of exposure to COVID-19 patients (from February 2021).

The first COVID-19 case was detected in the country during February, 28, 2020. According to epidemiological and, subsequently, molecular genetic data, the case is classified as imported (from Iran) [12].

Over the following 4 weeks (March 2-29, 2020), mainly sporadic cases were registered, mostly among individuals returning from Europe (Italy, Portugal) and those who had contact with individuals arriving from abroad.

In the initial phase of the epidemic, which lasted for 5 calendar weeks (calendar weeks 9-13, March 2-29, 2020), 1 to 49 COVID-19 cases, with an average weekly incidence rate of 0.2 cases per 100,000 population, were registered. Only one month after the first case was reported (March 30, 2020), 58 new cases per day were detected.

During calendar week 14 (March 30, 2020 – April, 5, 2020), local coronavirus spread was noted and the first COVID-19 wave began.

The maximum incidence rate was recorded during calendar week 20 (May, 11-17, 2020) and amounted to 71.4 cases per 100,000 population. The incidence rate peaked on May 17, 2020, when 969 cases were reported.

Thereafter, there was a gradual decrease in the number of cases with a minimum incidence level during calendar weeks 32-33 (9.3 and 6.2 cases per 100,000 population, respectively).

Overall, the first rising incidence period continued in the country for 25 weeks (calendar week 9-33, February, 28, 2020 -August, 16, 2020). The total number of cases was 69,516, with an average weekly incidence rate of 29.7 cases per 100,000 population (Figure 1).

**Figure. 1.**
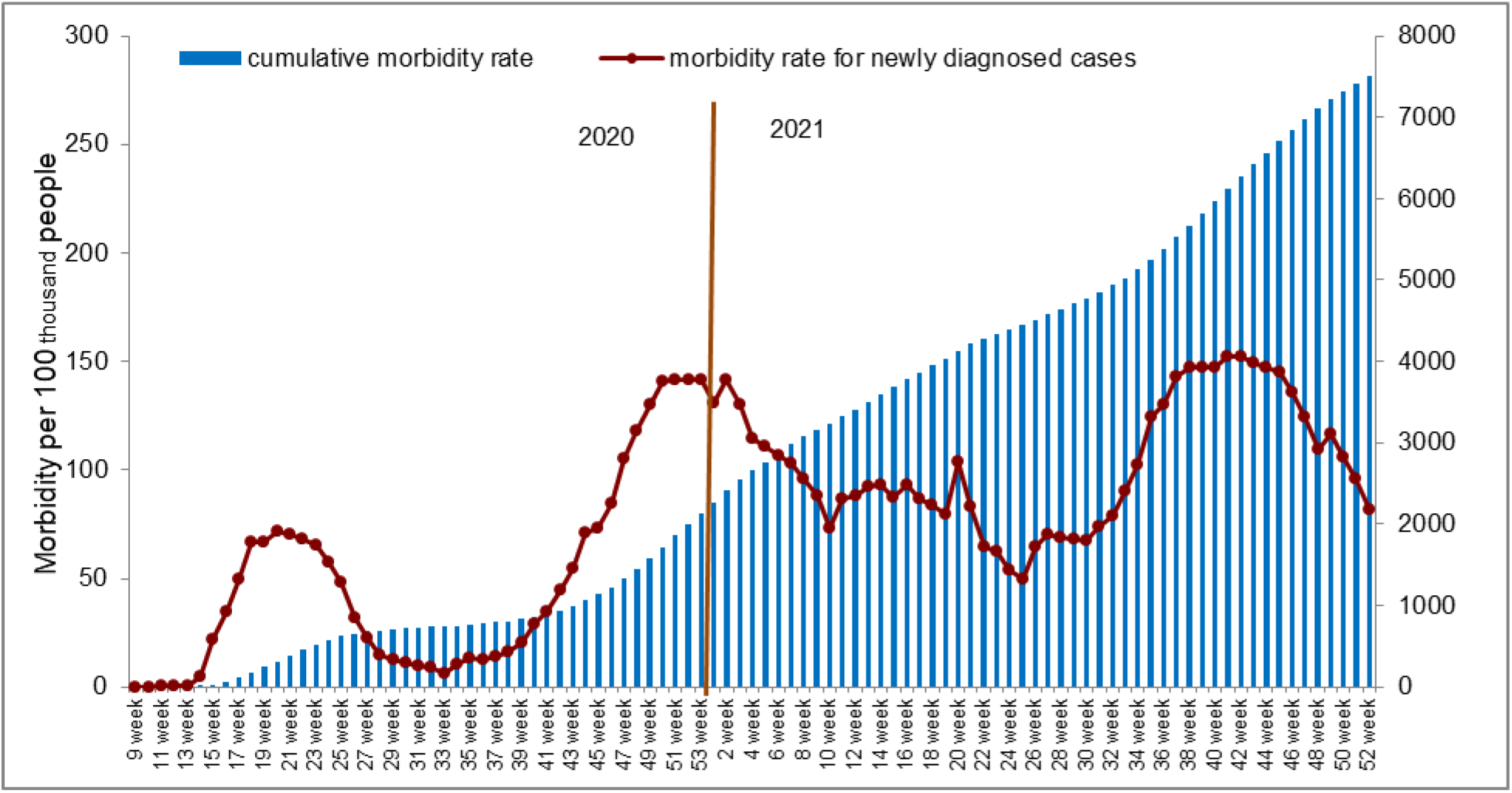
Dynamics of the incidence of COVID-19 in the population of the Republic of Belarus during in 2020-2022 years.

As of calendar week 34, an upward trend began with a rapid increase in the number of cases. The maximum incidence rate was recorded on calendar week 2, 2021 (January, 11-17, 2021): 141.8 cases per 100,000 population and was 1.98 times higher than the maximum rate of the first rising incidence period. The peak incidence was recorded on January 13, 2021: 1,972 COVID-19 cases, which is twice higher than the previous peak incidence.

Overall, the second, more intense and prolonged, rising incidence period lasted for 30 calendar weeks: until calendar week 10, 2021 (March 08-14, 2021). The average weekly incidence rate increased 2.8 times compared to the first rising incidence period and amounted to 83 cases per 100,000 population. During this period, COVID-19 cases were noted for which no epidemiological link could be established between patients (i.e. patients had no history of contact with COVID-19 patients or other respiratory infections).

Molecular genetic analysis of SARS-CoV-2 isolates collected in Belarus from February 2020 to February 2021 showed the spread of 4 COVID-19 clades in the country: O (5%), G (25%), GR (27.5%), GH (40%) and GRY (2.5%). A wide representation of the Pangolin lineage (52.5% of the genomes belonged to lineage B.1.) indicates that the SARS-Cov-2 virus came to Belarus from various areas [13].

During the next 4 calendar weeks (March 15, 2021 to April 11, 2021), another upward trend was observed, with daily cases ranging from 876 to 1,651. During calendar weeks 15-25, COVID-19 incidence showed a decreasing trend. Alpha variant (B.1.1.7) was dominating. It was identified on February 27, 2021.

As of calendar week 26, 2021, another rise in the incidence was recorded, due to the spread of the Delta SARS-CoV-2 variant in Belarus. Delta variant of SARS-CoV-2 has since become the prevalent in the country. At the end of December 2021, the first cases of infection of the Omicron SARS-CoV-2 variant were identified in the Republic of Belarus (Figure 2).

**Figure 2.**
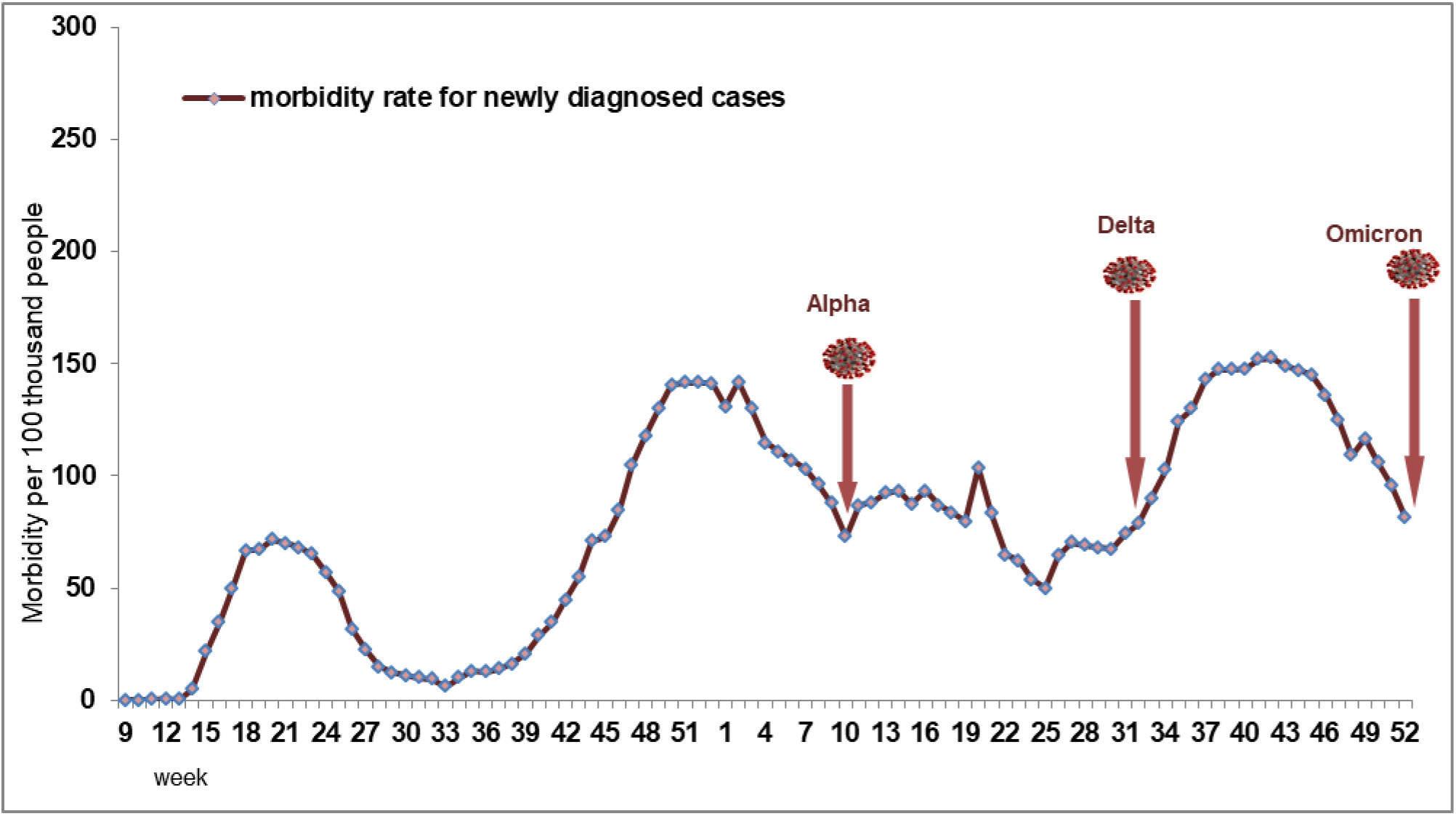
Results of molecular genetic analysis of SARS-CoV-2 isolates collected in Belarus

A total of 701,699 COVID-19 cases were reported from February 28, 2020 to January, 3, 2022. The cumulative incidence rate was 7,905.1 per 100,000 population.

The largest number of cases of the disease was registered among the adult population (90%). The proportion of children in the structure of cases -10% (Figure 3).

**Figure 3.**
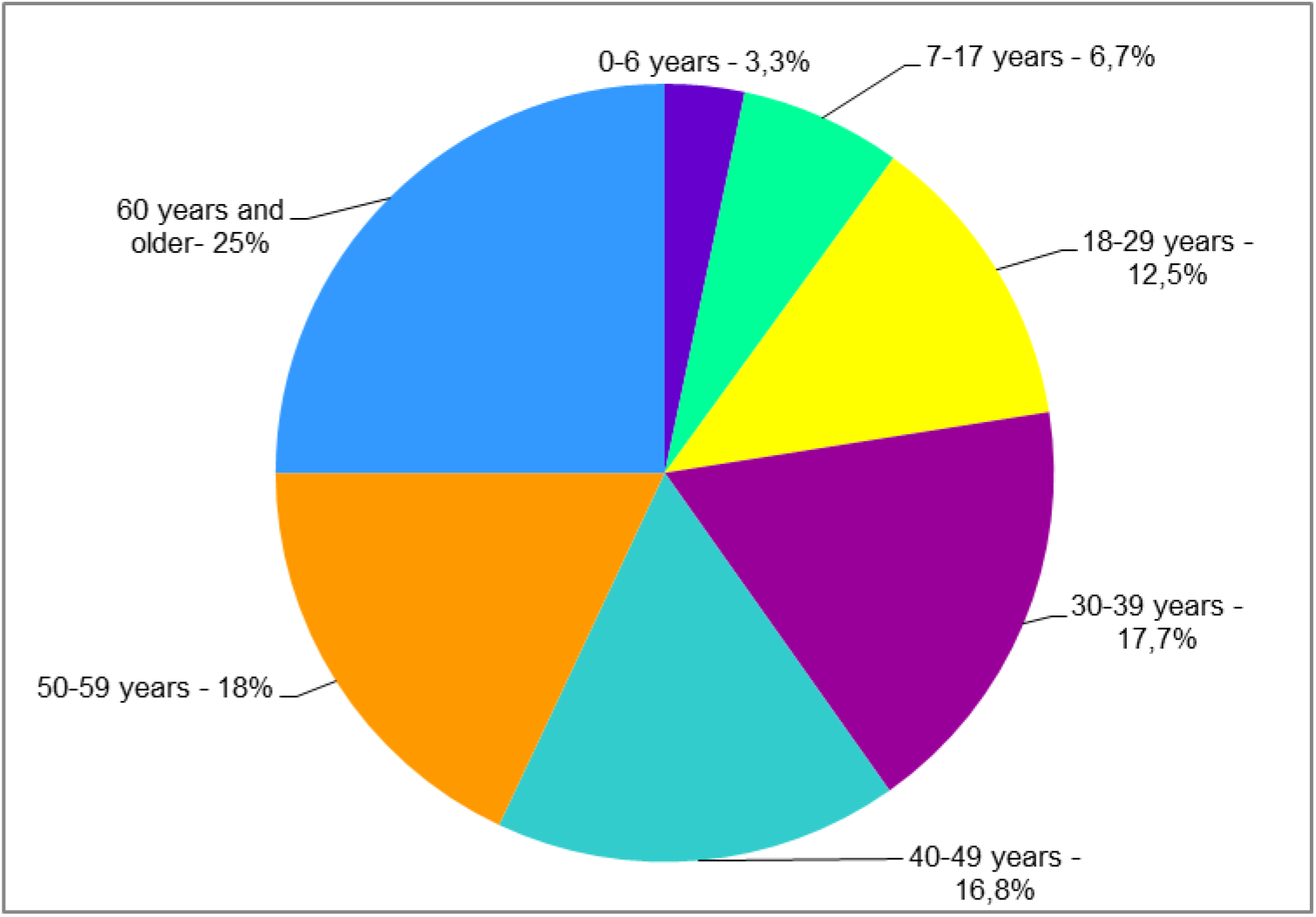
Age structure of COVID-19 cases in the Republic of Belarus in 2020-2022

The first COVID-19 fatality was registered on March 31, 2020, 6 weeks after the first case was detected. As of January, 3, 2022, 5 624 patients, or 0.8% of the total number of cases, died.

An important indicator of the COVID-19 pandemic development intensity is the infection spread index (R).

At the initial stage of the COVID-19 pandemic, the infection spread index (R) was characterized by marked variability (from 0.3 to 26), which probably determined a rapid infection spread.

During the first rising incidence period, R ranged from 0.7 to 1.6; during the second one, R ranged from 0.8 to 1.3. From August of this year until mid-September, R was more than 1 (with a maximum level of 1.3). Further, for 8 weeks (20.09.2021-14.11.2021), R remained at level 1. And only from the 46th calendar week the distribution index was less than 1.

### 3.2. Characteristics of sanitary and anti-epidemic measures

In the Republic of Belarus, organizational work to prevent the occurrence and minimize the risk of the spread of COVID-19 was launched in January 2020 on the basis of monitoring the epidemiological situation in the world, the recommendations of the World Health Organization and taking into account the available data on the epidemiology of this infection. In particular, additional measures have been introduced to strengthen sanitary and quarantine control at checkpoints across the State border of the Republic of Belarus in respect of arriving citizens, and laboratory examination has been organized. The development and implementation of the Comprehensive Action Plan to minimize the risk of importation and spread of a new coronavirus infection was ensured, taking into account various options for the development of the epidemic process in the country, providing for the interaction of interested ministries and departments.

Since the registration of the first cases of the disease, the sanitary and epidemiological service of the republic has carried out an epidemiological investigation, identifying contact persons, organizing the necessary sanitary and anti-epidemic measures at the place of residence (study, work, temporary stay).

Particular attention in the framework of countering the spread of COVID-19 is paid to compliance with sanitary and epidemiological requirements in health organizations. Relevant legal acts and recommendations on the organization of the functioning of outpatient polyclinic and inpatient healthcare organizations in the conditions of COVID-19 registration, educational and methodological films on the procedure for using protective clothing and respiratory protection by employees have been developed.

Taking into account scientific data and practical experience, recommendations have been developed for carrying out sanitary and anti-epidemic measures in the implementation of various activities (educational institutions, health and sanitary-resort institutions, catering and trade facilities, sports, transport, tourist organizations and others), based on the basic principles of disease prevention: social distancing, including minimization of contacts, use of respiratory protection, compliance with the rules of personal hygiene and other measures.

As of December 2020, vaccination has been deployed nationwide: health workers, as the top occupational risk group, were the first to be vaccinated.

The National Plan on Vaccination against COVID-19 for 2021-2022 approved by the government of the Republic of Belarus defines immunization tactics including 4 stages of sequential inclusion of specific groups in the vaccination campaign covering 60% of the population.

The first 3 stages provide for immunization of individuals in various risk groups: healthcare and pharmaceutical workers; social care and educating institution staff; individuals aged 61 and older and those with chronic diseases; other individuals with a higher risk of infection and a more serious course of the disease. Then the rest of the adult population are to be vaccinated.

### 3.3. Assessment of the safety (tolerance) of the vaccines used and the epidemiological efficacy of the vaccination

For immunization in the Republic of Belarus, the following are used: the vector vaccine Gam-COVID-Vac or Sputnik V (Russian Federation), the inactivated vaccine against SARS-CoV-2 (Vero Cell) Sinopharm / BIBP (China), as well as from September 2021 -the Sputnik Light vaccine. Vaccination was carried out in accordance with the instruction for medical use.

As of January 3, 2022, in the Republic of Belarus, 3,732,257 (39.9%) residents were vaccinated with the first dose of the vaccine. Taking into account the one-time immunization with the use of the Sputnik Light vaccine, the coverage of the full course of preventive vaccinations in the population of the country was 31.8%.

Full vaccination against COVID-19 was provided to 199 195 (91,2 %) of healthcare workers; 24 162 (81,7 %) of workers in 24-hour institutions; 213 501 (70,5 %) of workers in educational institutions; 1 329 511 (56,3 %) of individuals aged over 60 and those with chronic diseases.

The results of satellite V, Sinopharm vaccines for the period January-November 2021 showed that drugs were well tolerated: vaccinated reported 125,500 adverse reactions (1.89% (95% CI 1.8-1.9) of the number of doses administered). In terms of severity, adverse reactions were mild (91,9% (95% ДИ 91.7-92.0)) and moderate (8,1% (95% ДИ 7.9-8.3). The most frequent adverse reactions included injection place pain, hyperthermia and asthenia.

In 4 vaccinated (persons aged 30-35, 40-45, 50-54 years), serious adverse reactions were registered (2 – after the introduction of component I of the Sputnik V vaccine: “Allergic urticaria to the introduction of ILS” (ICD code 10 - L 50.0) and “Allergic reaction of the anaphylactoid type to the introduction of the Gam-COVID-Vac vaccine” (ICD code 10 - T.88.6); 1 – after the introduction of component II of the Sputnik V vaccine: “Allergic urticaria to the introduction of ILS” (ICD code 10 - L 50.0); 1 – after the introduction of an inactivated vaccine against SARS-CoV-2 (Vero Cell) Sinopharm / BIBP (PRC): “Allergic urticaria to the introduction of ILS”. All patients fully recovered without consequences.

According to the research, these reactions were caused by individual reactions to an adequately prescribed and correctly administered medicinal product.

Based on the calculated indicators of the incidence of vaccinated and unvaccinated, the coefficient and index of the epidemiological effectiveness of vaccination were calculated.

The epidemiological efficacy rate was 89,8%, the epidemiological efficacy index was 9,8. Thus, the results obtained indicate a high preventive efficacy of vaccines against COVID-19 used in the country.

## 4. Discussion

The emergence and active spread of COVID-19 has required countries to respond quickly. At the initial stage, the main direction of preventing the spread of infection and protecting the population in the Republic of Belarus, as in other countries [14], was to carry out sanitary and anti-epidemic measures of the country’s territory. The work carried out made it possible to postpone the emergence and spread of infection in the republic to a certain extent: the first case of COVID-19 was detected in Minsk at the end of February 2020.

Taking into account the development of the epidemiological situation, certain sanitary and anti-epidemic measures were introduced (in particular, isolation of patients and contact persons, measures to minimize the risk of the spread of COVID-19 in various collectives (educational institutions, health organizations and others).

The country’s response to COVID-19, reflecting the implementation of a proactive response strategy, has helped to contain the spread of infection without declaring a lockdown.

According to the WHO classification [15], the global epidemic process of COVID-19 in the world is characterized by 4 stages of development – from no cases in early 2020 to detected cases where most individuals had no history of contact with COVID-19 patients. The above stages of the development of the epidemic process can be traced in the Republic of Belarus.

One of the most important strategic preventive measures was the vaccination of the population. According to a systematic review of 11 studies, most of the reactions that occurred after the introduction of COVID-19 vaccines were mild to moderate and resolved within 3–4 days [16]. The most common local reactions included pain at the injection site, swelling and redness, systemic reactions including fever, fatigue, myalgia and headache.

Vaccines against COVID-19 (Gam-COVID-Vac” (Russia), inactivated vaccine against SARS-CoV-2 (Vero Cell) Sinopharm / BIBP (China) demonstrated a high safety profile in mass vaccination of the population of the Republic of Belarus. The identified adverse reactions were considered as mild to moderate and did not require medical intervention.

Serious adverse reactions to the introduction of vaccines were registered only in 4 vaccinated, their main cause is an allergy to the components of the vaccine.

Our findings indicate that both vaccine demonstrated its epidemiological efficacy. During the 11 months of follow-up, the incidence in the vaccinated was 86.6% lower than in the unvaccinated.

## Data Availability

All data produced in the present work are contained in the manuscript

